# Provision of professional interpreters and Heart School attendance for foreign-born compared with native-born myocardial infarction patients in Sweden

**DOI:** 10.1101/2023.09.12.23295463

**Authors:** Sammy Zwackman, Margret Leosdottir, Emil Hagström, Tomas Jernberg, Jan-Erik Karlsson, Sofia Sederholm Lawesson, Halldora Ögmundsdottir Michelsen, Annica Ravn-Fischer, John Wallert, Joakim Alfredsson

**Affiliations:** Department of Health, Medicine and Caring Sciences, Division of Diagnostics and Specialist Medicine, Unit of Cardiovascular Sciences, Linköping University, Linköping, Sweden; Department of Cardiology, Skane University Hospital, Malmö Sweden; Department of Clinical Sciences, Faculty of Medicine, Lund University, Malmö, Sweden; Department of Medical Sciences, Cardiology and Uppsala Clinical Research Centre, Uppsala University, Sweden; Department of Clinical Sciences, Danderyd Hospital, Karolinska Institute, Stockholm Sweden; Department of Internal Medicine, County Hospital Ryhov, Jönköping, Sweden; Institution of Medicine, Department of Molecular and Clinical Medicine, Sahlgrenska Academy, Gothenburg University. Department of Cardiology, Sahlgrenska University Hospital, Gothenburg, Sweden; Center for Psychiatry Research, Department of Clinical Neurosciences, Karolinska Institute, Stockholm Sweden and Stockholm HealthCare Services, Region Stockholm, Huddinge, Sweden; Institution where the study was performed: Department of Health, Medicine, and Caring Sciences, Division of Diagnostics and Specialist Medicine, Unite of Cardiovascular Science, Linköping University, Linköping, Sweden

**Keywords:** Myocardial infarction Secondary prevention Professional interpreters Language barriers Patient education

## Abstract

**Background and aims:** Interactive patient education, referred to as Heart School (HS), is an important part of cardiac rehabilitation (CR) after myocardial infarction (MI), which has been associated with improved outcomes. Little is known about HS attendance among foreign-born patients. The aims were to assess; 1) HS attendance in foreign-born versus native-born patients, 2) the association between the provision of professional interpreters and HS attendance, and 3) secondary prevention goal attainment after MI based on HS attendance.

**Methods:** The provision of professional interpreters during post-MI follow-up was assessed by a questionnaire sent to all 78 Swedish CR sites. Patient-specific data was retrieved from the SWEDEHEART registry. The association between provision of professional interpreters and HS attendance was estimated with logistic regression models. HS attendance and attainment of secondary prevention goals by country of birth were investigated.

**Results:** In total, 8377 patients <75 years (78% male) were included. Foreign-born (19.8%) had higher prevalence of cardiovascular risk factors and were less likely to attend HS (33.7 vs 51.3%, p<0.001), adjusted odds ratio (OR) 0.59 (95% confidence interval (CI) 0.52-0.68), compared with native-born patients. CR centers providing professional interpreters had higher HS attendance among foreign-born (adjusted OR 1.55, 95% CI 1.20-2.01) but not among native-born patients. Attending HS was similarly associated with improved secondary prevention goal attainment in both groups.

**Conclusion:** Despite similar positive association between HS attendance and attainment of secondary prevention goals, foreign-born patients attended HS less often. With provision of professional interpreters HS attendance appears to increase in foreign-born patients.

## Introduction

Cardiovascular (CV) disease is a major cause of death world-wide and myocardial infarction (MI) is the most frequent acute CV disease (1, 2). MI outcomes have improved during the last decades due to improvement in acute management, favorable lifestyle changes, and more effective primary and secondary preventive therapies (3–5). A majority of MI cases are attributed to modifiable risk factors and are largely preventable (3). Yet, the risk of recurrency remains high, and optimization of CV risk factors and lifestyle changes post-MI are of uttermost importance (6–8). Secondary prevention by cardiac rehabilitation (CR), including professional support to modify unfavorable lifestyle, improve drug adherence, provide patient education, and increase patient empowerment, has been shown to reduce the risk of recurrent CV events and death (5, 9). Accordingly, international guidelines have repeatedly advocated the use of CR post-MI (6, 10). In Sweden, Heart School (HS) is a core CR element providing interactive education on diet, exercise, smoking cessation, and health promotion in a group setting. Previous studies showed that attending HS was associated with lower risk of recurrent CV events and favorable long-term prognosis (9, 11, 12).

Many developed countries have gone through demographic changes because of immigration and influx of refugees (13, 14). In Sweden, the proportion of foreign-born residents increased from 11.7% in 2000 to 19.7% in 2020 (15). Consequently, language has become an increasingly important healthcare barrier. Previous studies from United States and Canada have shown that patients with limited English proficiency (LEP) had fewer physician visits and were less likely to receive preventive services (16–18). In Denmark, CR core components were provided to a lesser degree to non-Danish speaking patients without improvement over time (19, 20). The use of professional interpreters has been suggested to bridge language barriers and improve outcomes in a wide range of patient populations (18, 21–24). To our knowledge, the use of professional interpreters has not been studied in MI patients and no previous study has assessed the association between provision of professional interpreters and attendance to important CR elements in post-MI patients with limited majority language proficiency (the language predominantly spoken by healthcare professionals in the healthcare system that the patients attend).

The primary aim was to investigate HS attendance in foreign-born and native-born MI patients as well as the association between HS attendance and the provision of professional interpreters at CR follow-up visits. In addition, secondary prevention goal attainment based on HS attendance was evaluated.

## Methods

This was a sub-study to the Perfect Cardiac Rehabilitation (Perfect CR) study which has been previously described (25, 26). For Perfect CR, organizational and patient specific data was collected and merged into one database. Organizational variables were collected by a detailed questionnaire sent to all 78 CR centers in Sweden. These centers were actively reporting patient-level data to the Swedish Web-system for Enhancement and Development of Evidence-based care in Heart Disease Evaluated According to Recommended Therapies (SWEDEHEART). The purpose of Perfect CR was to assess key elements of guidelines-recommended CR, including both structure and processes applied in the CR programs and their outcomes (25, 27, 28). The survey gathered information on routine provision of professional interpreters to non-Swedish speaking patients during CR follow-up after MI. A professional interpreter is defined as someone who is specialized in interpreting from one spoken language to another and facilitating communication between a foreign-born patient with low Swedish proficiency and healthcare professionals. In Sweden, the information about patients’ native language and the need for interpreters is registered in patient records. CR follow-up visits and professional interpreters are booked in advance. In this study, if professional interpreters were not provided by CR centers, ad hoc interpreters such as family members and friends were allowed to interpret in certain centers. Ad hoc interpreters (family members and friends) were not the subject of this study. In this analysis, CR centers were defined as centers providing *professional* interpreters or not.

The study population consisted of all MI patients hospitalized in Sweden during the predefined study period reflected by the questionnaire (1^st^ Nov. 2015 until 31^st^ Oct. 2016) with one year follow-up. The inclusion criteria were: 1) type 1 MI diagnosis, 2) age between 18 and 74 years, and 3) attending at least one of the two CR visits during the first-year post-MI at which data is registered in SWEDEHEART. Patient specific variables were retrieved from the SWEDEHEART registry and included baseline characteristics, in-hospital management, and follow-up including risk factor management, (*Table 1, Table S1* and *S2*). The registry is regularly monitored by external monitors, with more than 95% agreement between registered information and medical records (29). Furthermore, census-based individual-level data including information about death, country of birth, marital status, employment, education level, and disposable income were retrieved from Statistics Sweden, which is the government agency responsible for providing official statistics (30).

**Table 1.** Baseline characteristics.

| Table 1. Baseline characteristics |  |  |  |
| --- | --- | --- | --- |
|  | Foreign-born<br>N=1655 | Native-born<br>N=6722 | P-value |
| Age (SD) | 59.5 (9.3) | 63.4 (8.3) | <0.001 |
| Female sex | 364 (22.0) | 1701 (25.3) | 0.01 |
| Risk factors and comorbidity |  |  |  |
| Smoking status |  |  |  |
| Current smoker | 713 (43.3) | 1751 (26.2) | <0.001 |
| Previous smoker | 556 (33.8) | 2654 (39.7) |  |
| Never smoked | 377 (22.9) | 2287 (34.2) |  |
| Hypertension | 750 (45.6) | 3219 (48.1) | 0.07 |
| Diabetes mellitus | 477 (28.9) | 1456 (21.7) | <0.001 |
| Previous MI | 382 (23.1) | 1273 (19.0) | <0.001 |
| Previous PCI | 345 (21.0) | 1110 (16.6) | < 0.001 |
| Previous CABG | 107 (6.5) | 379 (5.7) | 0.20 |
| Previous revascularization | 385 (23.5) | 1270 (19.0) | < 0.001 |
| Previous stroke | 70 (4.2) | 296 (4.4) | 0.84 |
| Known LVD | 96 (5.9) | 341(5.2) | 0.24 |
| Medication at admission |  |  |  |
| Aspirin | 429 (26.5) | 1609 (24.3) | 0.07 |
| P2Y12-inhibitor | 83 (5.1) | 261 (3.9) | 0.04 |
| ACE-inhibitor | 351 (21.7) | 1237 (18.7) | 0.01 |
| ARB | 221 (13.7) | 1268 (19.2) | <0.001 |
| Oral anticoagulation | 64 (3.9) | 310(4.7) | 0.23 |
| Betablockers | 491 (30.4) | 1865 (28.2) | 0.09 |
| Statin | 488 (30.1) | 1814 (27.4) | 0.03 |
| Insulin | 159 (9.8) | 627 (9.5) | 0.67 |
| Oral diabetes medication | 291 (17.9) | 796 (12.0) | <0.01 |
| Socioeconomic factors |  |  |  |
| Marital status |  |  |  |
| Married /living together | 980 (59.3) | 3834 (57.1) | 0.11 |
| Living alone | 674 (40.7) | 2885 (42.9) |  |
| Income (quintiles) |  |  |  |
| 1 | 629 (38.0) | 982 (14.6) | <0.001 |
| 2 | 312 (18.9) | 1339 (19.9) |  |
| 3 | 281 (17.0) | 1384 (20.6) |  |
| 4 | 240 (14.5) | 1477 (22.0) |  |
| 5 | 192 (11.6) | 1535 (22.8) |  |
| Education |  |  |  |
| Less than 10 years | 496 (31.3) | 1814 (27.0) | <0.001 |
| 10-12 years | 684 (43.2) | 3385 (50.5) |  |
| College/university level | 405 (25.6) | 1510 (22.5) |  |
| Clinical and laboratory findings at admission |  |  |  |
| Systolic BP mmHg (SD) | 149.4 (28.2) | 150.9 (27.7) | 0.02 |
| Cholesterol mmol/l (SD) | 5.0 (1.3) | 5.0 (1.3) | 0.33 |
| LDL-cholesterol mmol/l (SD) | 3.1 (1.2) | 3.1 (1.1) | 0.89 |
| eGFR ml/min (SD) | 90.5 (28.4) | 87.5 (24.5) | <0.001 |
| BMI (SD) | 28.4 (4.7) | 27.9 (4.7) | <0.001 |
| Type of MI |  |  |  |
| STEMI | 642 (38.8) | 2608 (38.8) | 1.00 |
| LV function during index |  |  |  |
| LVEF ≥50% | 1014 (67.5) | 3932 (64.9) | 0.01 |
| LVEF = 40-49% | 293 (19.5) | 1292 (21.3) |  |
| LVEF = 30-39% | 142 (9.5) | 590 (9.7) |  |
| LVEF <30 | 51 (3.4) | 191 (3.2) |  |
| **Medication at discharge** |  |  |  |
| Aspirin | 1595 (96.4) | 6414 (95.5) | 0.09 |
| P2Y12-inhibitors | 1506 (91.0) | 6166 (91.7) | 0.32 |
| Dual anti-platelet therapy | 1459 (88.2) | 5926 (88.2) | 0.97 |
| Oral anti-coagulation | 137 (8.3) | 618 (9.2) | 0.25 |
| ACE-inhibitors | 1100 (66.5) | 4188 (62.3) | 0.01 |
| ARB | 294 (17.8) | 1627 (24.2) | < 0.001 |
| Betablockers | 1510 (91.2) | 6027 (89.7) | 0.06 |
| Statins | 1605 (97.0) | 6534 (97.2) | 0.56 |
| Insulin | 190 (11.5) | 649 (9.7) | 0.03 |
| Oral diabetes medication | 333 (20.1) | 864 (12.9) | <0.001 |
| Results are presented as numbers (percentages) and mean (SD). Abbreviation: SD, standard deviation; PCI, percutaneous coronary intervention; CABG, coronary artery bypass graft; LVD, left ventricular dysfunction; BMI, body mass index; MI, myocardial infarction; STEMI, ST-elevation myocardial infarction; LV, Left ventricle; LVEF, LV ejection fraction; ACE-inhibitors, angiotensin converting enzyme; ARB, angiotensin receptor blocker; BP, blood pressure; eGFR, estimated glomerular filtration rate; BMI, body mass index; STEMI, ST-elevation myocardial infarction. |  |  |  |

### Exposure and outcome definitions

In the primary analysis, HS attendance in foreign-born was compared with native-born (born in Sweden) patients. Secondly, the association between HS attendance (outcome) and the provision of professional interpreters at follow-up visits (exposure) was assessed. Finally, attainment of secondary prevention goals and HS attendance in foreign-born and native-born patients (exposure) was investigated: 1) LDL-cholesterol <1.8 mmol (treatment goal during the study period), 2) systolic blood pressure <140mmHg, 3) attending physical training-based CR (supervised physical training at the hospital during the follow-up phase), and 4) smoking cessation. To achieve abstinence from smoking, current smokers were offered smoking cessation counselling by specially trained counsellors.

### Statistics

Continuous variables are presented as means with standard deviations (SD) and categorical variables as counts with percentages. Baseline and peri-procedural characteristics were compared based on if the patient was native-born or not. The Kolmogorov-Smirnov’s normality test was used to test whether data were normally distributed or not. The chi-square test was used for categorical variables and Student’s T-test or Mann-Whitney U test (depending on if the variable had a normal distribution or not) for continuous variables. For HS attendance, logistic regression models presenting crude and adjusted odds ratios (OR) with 95% confidence intervals (CI) were developed. Adjustments in the first model included age and sex; the second model added comorbidities, medications, and management variables (hypertension, smoking status, diabetes mellitus, previous MI (before index), previous revascularization, type of MI (ST-elevation MI [STEMI]/non STEMI,) and discharge medications (aspirin, P2Y12-inhibitors, beta-blockers, angiotensin-converting enzyme [ACE] inhibitors, angiotensin receptor blockers [ARB], statins, and diabetes medications)). Finally, in the third model socioeconomic variables were added (disposable income, education level and marital status).

For patient-related comparisons, native-born patients were the reference group, and for system-related comparisons, CR sites not offering professional interpreters were the reference group.

We performed two interaction tests: 1) country of birth (native-born or foreign-born) and the provision of professional interpreters on HS attendance. 2) country of birth (native-born or foreign-born) and HS attendance on secondary prevention goals.

A p-value <0.05 was considered statistically significant.

All analyses were performed using SPSS 29.0 statistical software package (SPSS Inc., Chicago, Illinois, USA).

### Ethical considerations

This study was performed in accordance with the Declaration of Helsinki and was approved by the Ethics Committee at Lund University (ethical permit number 2018-55).

In accordance with Swedish legislation, patients included in the SWEDEHEART registry were informed, before inclusion, about their participation in the registry and the right to opt out.

## Results

The study population consisted of 8,377 patients, out of which 1,655 (19.8%) were foreign-born. Foreign-born patients were more likely to be male (78.0 vs 74.7%), younger (mean age 59.5 vs 63.4 years), smokers (43.3 vs 26.2%), and to have diabetes (28.9 vs 21.7%), previous MI (23.1 vs 19.0 %), and previous revascularization with percutaneous coronary intervention (PCI) (21.0 vs 16.6%). Furthermore, foreign-born patients were overrepresented in the lower income classes (56.9 vs 35.5%) and higher education classes (25.6 vs 22.5%) (*Table 1*).

Compared with native-born, foreign-born patients were less likely to attend HS (33.7 vs 51.3%, p<0.001) corresponding to a crude OR of 0.48 (95% CI 0.43-0.54) (*Table 2*). After adjustment, the difference was moderately attenuated, especially after adding socioeconomic factors, but still significant (OR 0.59, 95% CI 0.52-0.68), (*Table 2*).

**Table 2.**
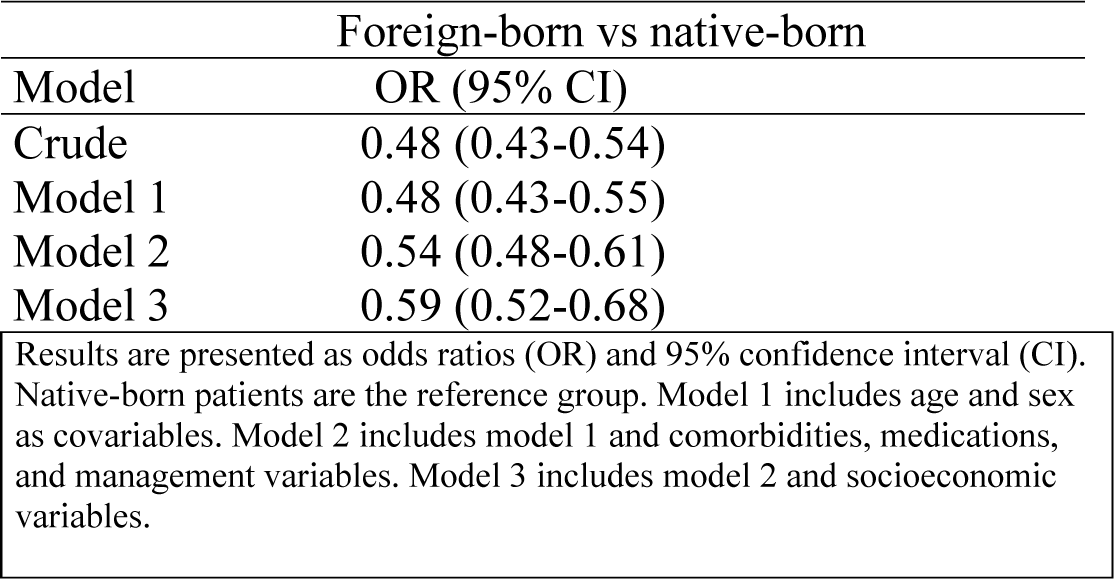
Heart School attendance in foreign-born patients.

| Foreign-born vs native-born |  |
| --- | --- |
| Model | OR (95% CI) |
| Crude | 0.48 (0.43-0.54) |
| Model 1 | 0.48 (0.43-0.55) |
| Model 2 | 0.54 (0.48-0.61) |
| Model 3 | 0.59 (0.52-0.68) |
| Results are presented as odds ratios (OR) and 95% confidence interval (CI). Native-born patients are the reference group. Model 1 includes age and sex as covariables. Model 2 includes model 1 and comorbidities, medications, and management variables. Model 3 includes model 2 and socioeconomic variables. |  |

Stratified analysis based on HS attendance showed minor differences in baseline characteristics in foreign-born patients. In contrast, native-born patients attending HS were less likely to be smokers, to have hypertension, diabetes mellitus, or previous revascularization, and they were more likely to be married/cohabiting, and to have higher education and disposable income (*Table S2*).

About one third of patients (33.2% of foreign-born and 31.9% of native-born patients, p<0.33) were followed at CR centers not routinely providing professional interpreters. A higher proportion of foreign-born patients attended HS (36.4 vs 27.5%, p=0.002, [OR 1.47, 95% CI 1.16-1.88]) at centers providing professional interpreters, whereas no differences were observed for native-born patients (50.5 vs 51.7%, p=0.39, [OR 1.05, 95% CI 0.94-1.17]) with a significant interaction test, p=0.001 (*Figure 1 and Table 3*).

**Figure 1.**
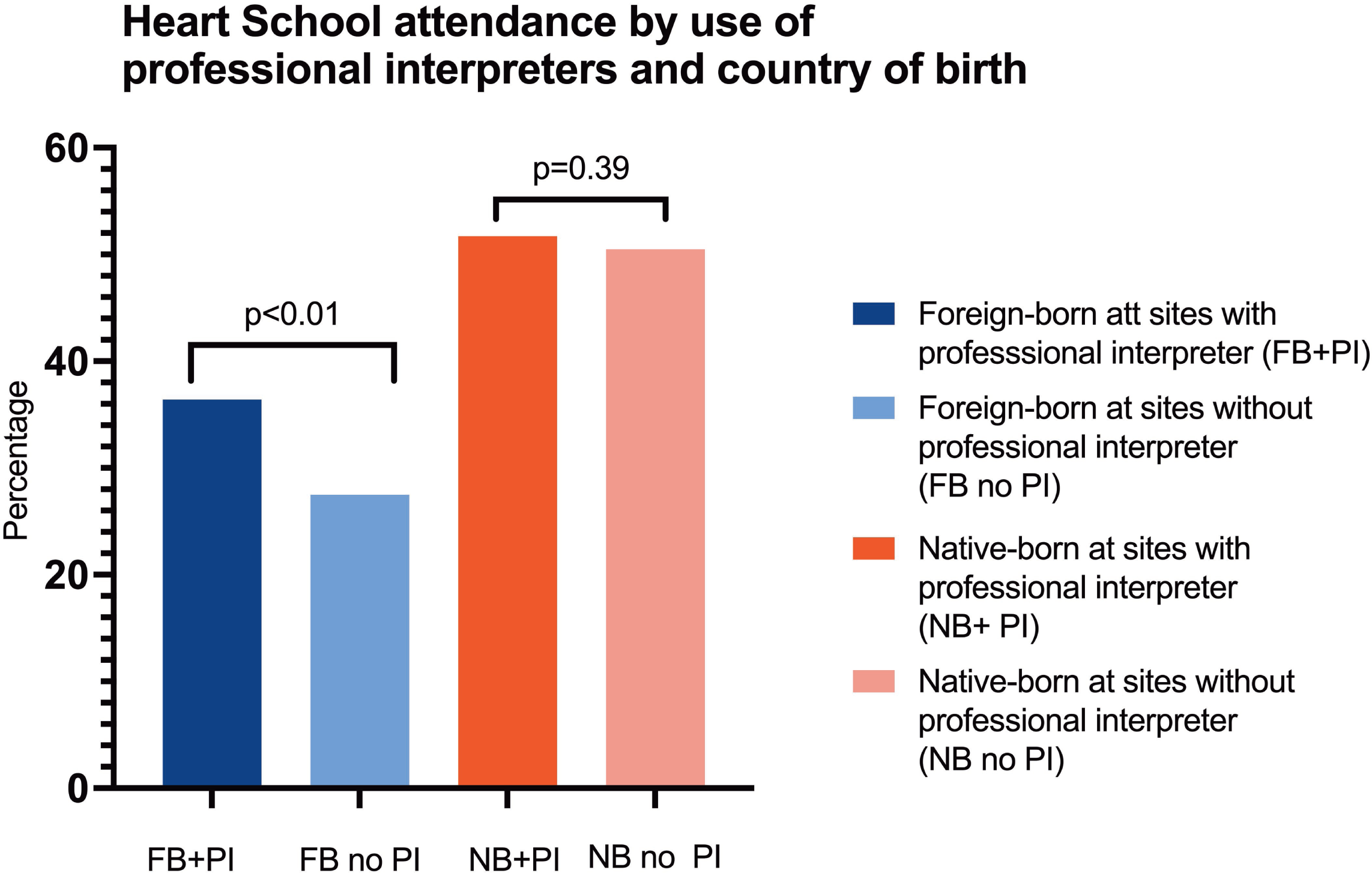
Heart school attendance after a myocardial infarction based on country of birth and sites offering professional interpreters or not. Abbreviations: FB, foreign-born; PI, professional interpreter; NB, native-born.

**Table 3.** Heart School attendance and provision of professional interpreters.

|  | Foreign-born | Native-born |
| --- | --- | --- |
| Model | OR (95% CI) | OR (95% CI) |
| Crude | 1.47 (1.16-1.88) | 1.05 (0.94-1.17) |
| Model 1 | 1.47 (1.16-1.88) | 1.05 (0.94-1.17) |
| Model 2 | 1.49 (1.17-1.90) | 1.08 (0.97-1.21) |
| Model 3 | 1.55 (1.20-2.01) | 1.08 (0.97-1.21) |

Results are presented as odds ratios (OR) and 95% confidence interval (CI). Sites not offering professional interpreters are the reference group. Model 1 includes age and sex as covariables. Model 2 includes model 1 and comorbidities, medications, and management variables. Model 3 includes model 2 and socioeconomic variables.

In foreign-born patients, neither adjustments for age and sex (model 1) (OR 1.47, 95% CI 1.16-1.88), nor model 1 plus comorbidities, medications, and management (OR 1.49, 95% CI 1.17-1.90) nor model 2 plus socioeconomic variables (OR 1.55, 95% CI 1.20-2.01), significantly changed the association between HS attendance and the routine provision of professional interpreters (*Figure 2A*). In native-born patients, there was no observed association before (OR 1.05, 95% CI 0.94-1.17), or after multivariable adjustment (OR 1.08, 95% CI 0.97-1.21) (*Figure 2B* and *Table 3)*.

**Figure 2A.**
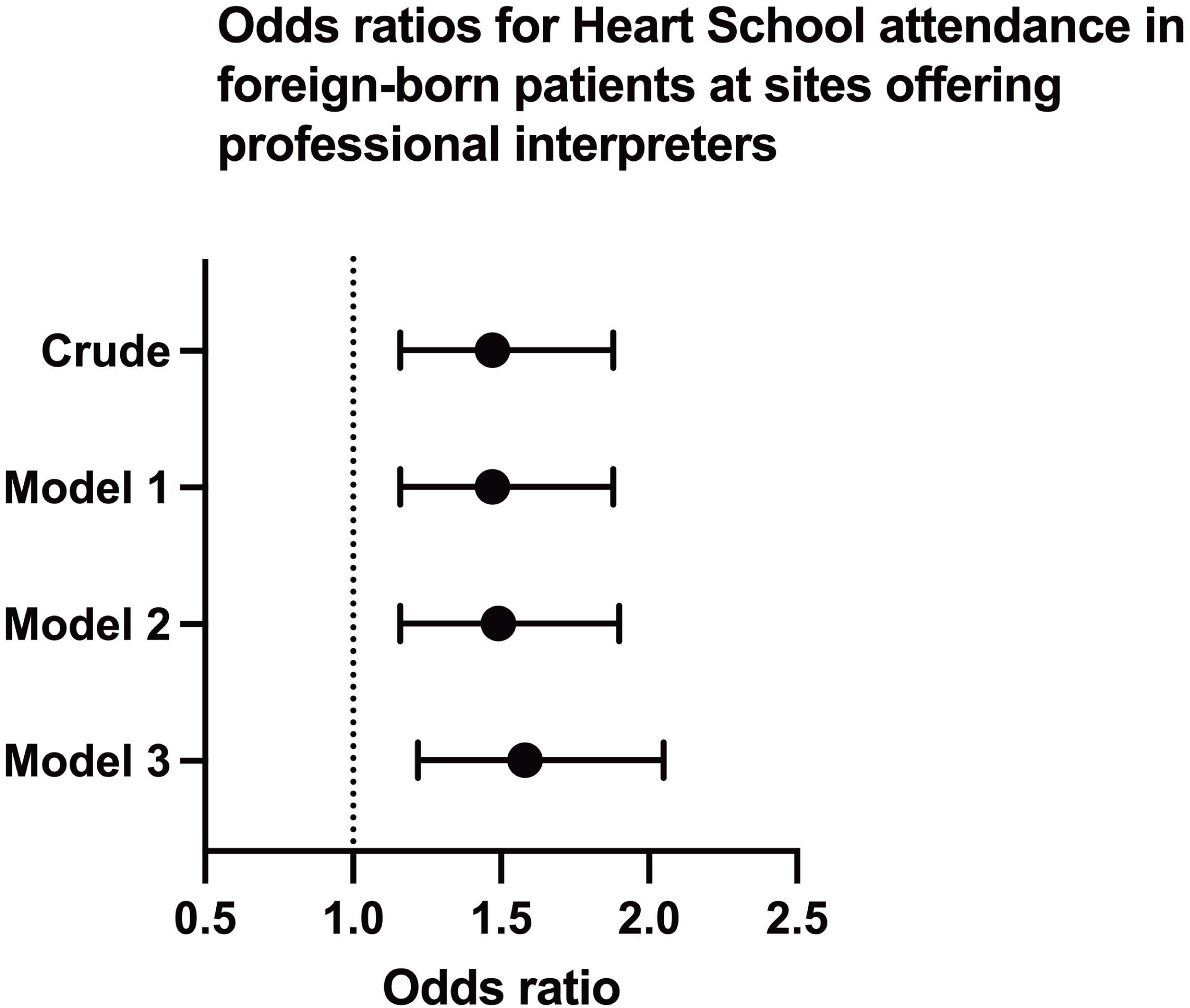
Odds ratios for participation in heart school for foreign-born patients at sites offering professional interpreters vs sites not offering professional interpreters. Model 1 includes age and sex as covariables. Model 2 includes model 1 and comorbidities, medications, and management variables. Model 3 includes model 2 and socioeconomic variables.

**Figure 2B.**
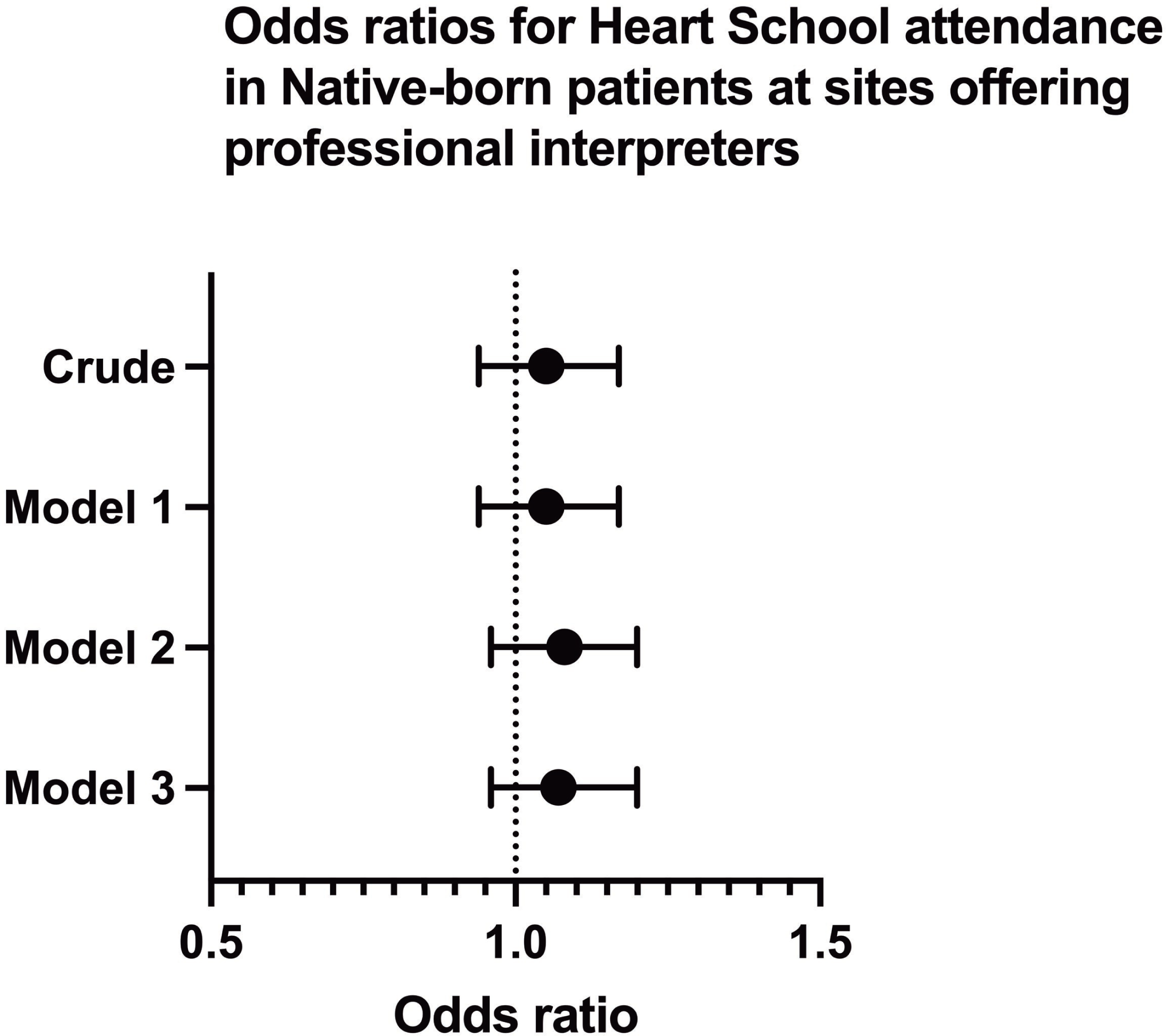
Odds ratios for participation in heart school for native-born patients at sites offering professional interpreters vs sites not offering professional interpreters. Sites not offering professional interpreters are the reference group. Model 1 includes age and sex as covariables. Model 2 includes model 1 and comorbidities, medications, and management variables. Model 3 includes model 2 and socioeconomic variables.

Attending HS was associated with better attainment of all four secondary prevention goals in foreign-born as well as native-born patients. In foreign-born patients, HS attendance was associated with a higher proportion of patients achieving LDL cholesterol <1.8 mmol/L (70.4 vs 64.3%, p=0.02), systolic blood pressure <140mmHg (91.0 vs 84.2%, p<0.001), attending physical training-based CR (71.2 vs 35.7% p<0.001), and smoking cessation after MI (68.3 vs 59.3%, p<0.001). In native-born patients, HS attendance was associated with achievement of all four treatment targets, LDL-cholesterol <1.8 mmol/L (69.4 vs 63.9%, p<0.001), systolic blood pressure <140mmHg (91.0 vs 84.2%, p<0.001), attending physical training-based CR (71.2 vs 35.7% p<0.001), and smoking cessation after MI (68.3 vs 59.3%, p<0.001). The interaction tests supported that the improved treatment goal achievements were similar in foreign-born and native-born patients, (*Table 4*). HS attenders had a higher participation in smoking cessation counselling compared to non-attenders, (32.9 vs 15.0%, p<0.001) in foreign-born and (23.1 vs 14.3%, p<0.001) in native-born patients *(Tables S3)*.

**Table 4.**
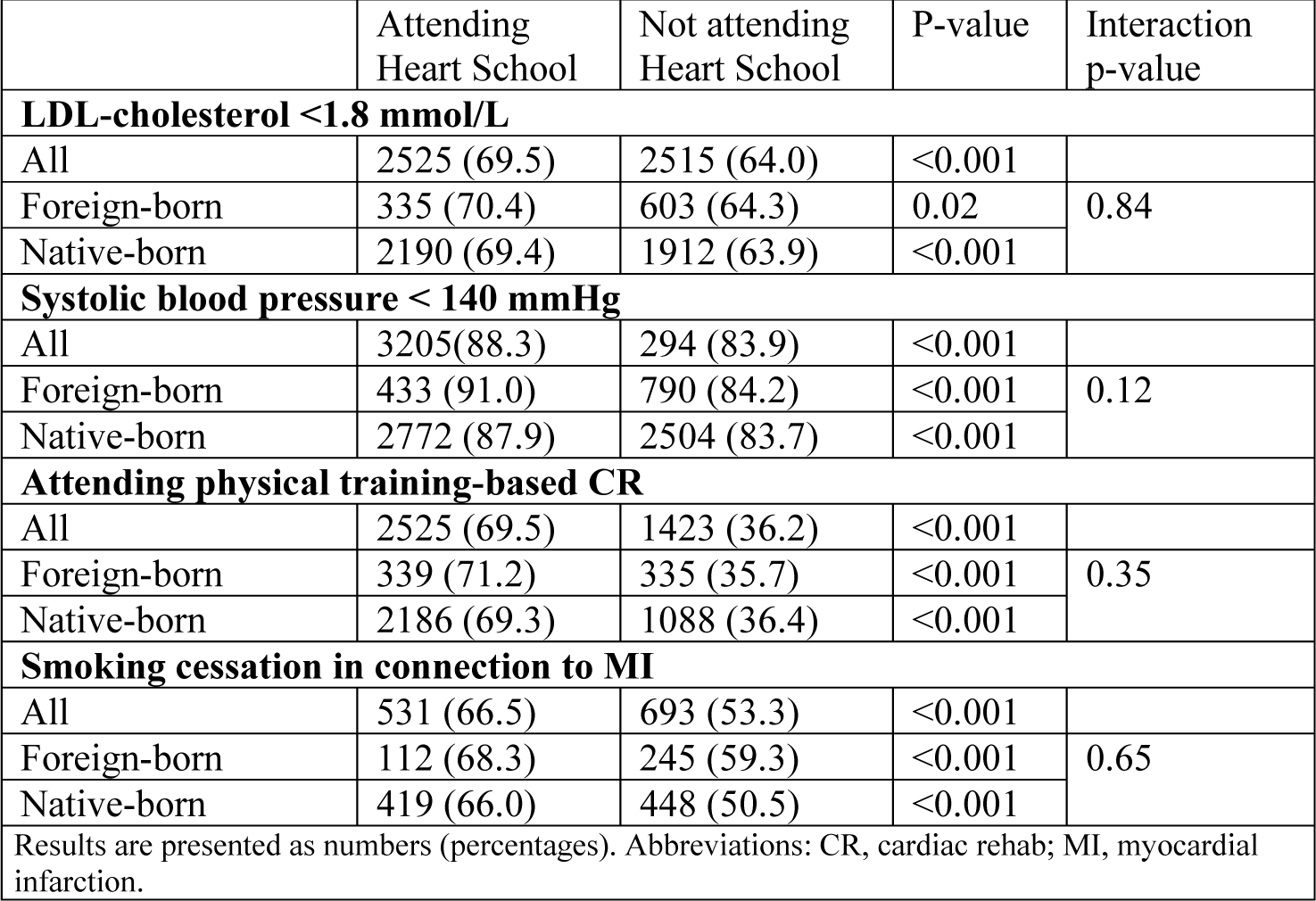
Attainment of secondary prevention goals in patients attending Heart School compared with patients not attending Heart School.

## Discussion

In the present study, routinely providing professional interpreters at the CR centers was associated with higher HS attendance among foreign-born but not in native-born patients, with a significant interaction test. HS was equally associated with better attainment of secondary prevention goals in foreign-born and native-born patients.

Foreign-born patients were more likely to present at younger age, were more often male and had a more severe CV risk profile, with higher prevalence of smoking, diabetes mellitus, previous MI and revascularization compared with native-born patients. These findings are congruent with previous studies, which have shown a different and more severe CV risk profile in foreign-born patients (31–34).

HS attendance was significantly lower among foreign-born compared with native-born patients. In English-speaking countries, previous studies have demonstrated that preventive services such as vaccination, disease screening, Pap tests, mammograms and physician visits are underutilized among patients with LEP which is in accordance with our results (16–18). The underutilization of preventive services among LEP patients has been attributed to socioeconomic factors, cultural aspects, and language barriers. In a Canadian study assessing the use of preventive services in English proficient and LEP patients, the association between language and preventive services persisted despite adjustment for socioeconomic factors and cultural aspects, reflecting the importance of language barriers in contact with health care (16, 17). This is in accordance with our results, showing very little impact of adjustment for socioeconomic factors on the likelihood of HS attendance for foreign-born patients. In studies from Denmark, the role of language barriers was suggested as the main reason for incomplete provision of core components of CR to non-Danish speaking MI patients. Furthermore, a lower uptake and a higher discontinuation of non-pharmacological prevention programs after MI (including physical training, dietary advice, and patient education) was reported in foreign-born compared with native-born patients, further supporting our results (19, 20). In the present study, routinely providing professional interpreters at the CR centers was associated with higher attendance in HS among foreign-born but not among native-born patients. A possible interpretation of these results is that the provision of professional interpreters bridges the language barrier between the healthcare professionals and foreign-born patients and improves HS attendance. It could be suggested that CR centers routinely providing professional interpreters may have more resources and better performance in general. However, the lack of association between centers providing professional interpreter and HS attendance in native-born patients further strengthens the interpretation that the provision of professional interpreters is not just a proxy for a well-functioning site but impacts HS attendance per se in foreign-born patients. Importantly, adjustment of potential confounders such as age, sex, comorbidities, medications, management, and socioeconomic variables did not change the association. Our findings indicate that the provision of professional interpreters may in fact help to overcome language barriers and improve CR attendance.

Attending HS or similar form of theoretically administered patient education after an MI have been associated with better attainment of secondary prevention goals and reduced CV event rates (9, 11, 12). A previous observational study showed that attending HS was associated with reduced CV and all-cause mortality (9). In a randomized trial, a relatively short structured educational programmed after MI was associated with lower risk of CV events (11). A meta-analysis investigating the length of patient education on attainment of secondary prevention goals, found that patient education was associated with better attainment of secondary prevention goals, irrespective of length of patient education (12). In accordance with previous findings, in our study, HS attendance was associated with better attainment of all four secondary prevention goals with no significant interaction between country of birth (native-born or foreign-born) and HS attendance on achievement of treatment goals.

To the best of our knowledge, the association between provision of professional interpreters and attendance to core components of CR (associated with attainment of secondary prevention goals) post-MI has not been studied previously. However, in a wide range of patient populations, the provision of professional interpreters improved outcomes by reducing unnecessary interventions in obstetric patients, lowering the rate of readmissions, and reducing the length of hospital stay in internal medicine patients (18, 21–23). Analogous to MI patients, in a previous study on LEP patients with diabetes and poor glycemic control, switching to a language-concordant physician to bridge the language barriers showed significant improvement both in glycemic control and LDL-cholesterol levels. (24). Taken together, these studies further emphasize the role of language barriers in healthcare and support our findings suggesting improved care in foreign-born patients with routine provision of professional interpreters.

### Strengths and limitations

The main strength of our study was the nation-wide inclusion of all 78 active CR centers in Sweden with 100% response rate addressing the use of professional interpreters, combined with high quality data from multiple registries for individual patients treated at these centers during the corresponding period. There are some important limitations to be mentioned. First, this was an observational study with its inherent limitations including the possibility of unmeasured confounding. However, multivariable adjustment did not significantly change the observed associations. Second, data on individual proficiency of foreign-born patients in the Swedish language was not available. Also, the provision of professional interpreters was reported at CR center level, not for individual patients. However, given the association observed for the whole foreign-born population, irrespective of individual need, it is likely that the association would be even stronger for patients with the lowest proficiency in the Swedish language.

## Conclusion

Foreign-born MI patients attended HS less often than native-born patients, despite similar association between HS attendance and attainment of secondary prevention goals in both groups. However, the provision of professional interpreters at follow-up visits at CR centers was associated with improved HS attendance among foreign-born patients. Therefore, provision of professional interpreters appears to improve CR among foreign-born patients.

## Funding

The Kamprad Family Foundation for Entrepreneurship, Research and Charity, The Swedish Research Council for Health, Working Life and Welfare, The Swedish Heart and Lung Association, The Swedish Heart and Lung Patient Organization, and The Swedish Cardiology Society, supported the work with unrestricted financial grants. The grant providers did not have any influence on study design, data interpretation or writing of the manuscript.

## Data Availability

SWEDEHEART does not allow individual data sharing to third party. Access to aggregated data might be granted following review by the SWEDEHEART steering committee. Such requests can be submitted to the SWEDEHEART steering committee for consideration.

## Acknowledgements

We gratefully acknowledge the support of the Kamprad Family Foundation for Entrepreneurship, Research and Charity in funding this project. We also thank The Swedish Research Council for Health, Working Life and Welfare, The Swedish Heart and Lung Association, The Swedish Heart and Lung Patient Organization and The Swedish Cardiology Society for their unrestricted grants. Without their generous grant, this study would have not been possible.

## Conflict of interests

Sammy Zwackman, Jan-Erik Karlsson, Halldora Ögmundsdottir Michelsen, Tomas Jernberg declared no conflict of interests. Margret Leosdottir received research grant from NovoNordisk and Pfizer and payments for lectures and educational events from AstraZeneca, NovoNordisk, Amgen, Sanofi and Amarin. Emil Hagström received research grants to institution from Pfizer and Amgen and a small personal fee from Amgen, NovoNordisk, Bayer and AstraZeneca for lectures and presentations. Annica Ravn-Fischer received payment for lectures and expert testimony from Amarin, Amgen, AstraZeneca, Boehringer, BMS, Novartis, NovoNordisk, Sanofi, Orion Pharma, Pfizer and Sanofi. Joakim Alfredsson received lecture fee from Boehringer Ingelheim, AstraZeneca, MSD, Bayer and Novartis and advisory board reimbursement from AstraZeneca and Novartis.

## Notes

### Competing Interest Statement

The authors have declared no competing interest.

### Clinical Trial

NA

